# Combining Transcranial Magnetic Stimulation with Antidepressants: A Systematic Review and Meta-Analysis

**DOI:** 10.1101/2022.11.03.22281857

**Authors:** Gopalkumar Rakesh, Patrick Cordero, Rebika Khanal, Seth S. Himelhoch, Craig R. Rush

## Abstract

Major depressive disorder (MDD) imposes significant disability on patients. In addition to antidepressants, brain stimulation modalities such as electroconvulsive therapy (ECT) and transcranial magnetic stimulation (TMS) have been helpful in treatment of MDD. Novel TMS paradigms like theta burst stimulation (TBS) have rapidly become popular due to their effectiveness.

Given that both antidepressants and TMS are commonly used together and affect neuroplasticity, we reviewed studies that administered both these as treatments for MDD. Unlike ECT wherein previous trials have shown that continuing pharmacotherapy is useful while giving ECT, there are no consensus guidelines on what to do with antidepressants when starting TMS. So, we reviewed two groups of studies – 1) those that administered TMS and antidepressant pharmacotherapy concurrently and 2) those wherein TMS augmented antidepressants or were an adjunctive intervention to antidepressants. We performed a meta-analysis for randomized clinical trials (RCTs) that administered TMS and antidepressants concurrently.

We found ten RCTs fulfilling criteria 1 and compared uniformly titrated antidepressant regimens combined with active versus sham TMS. We also found twenty studies fulfilling criterion 2, that used TMS as an augmenting or adjunctive intervention. Both groups of studies showed TMS combined with antidepressants had greater efficacy for treatment of MDD. We advocate for laboratory studies examining the interaction between TMS and antidepressants in a parametric fashion; in addition to randomized controlled trials that examine this combination to expedite remission in MDD.

## Introduction

Major depressive disorder (MDD) imposes significant disability and economic burden on patients^1^. Antidepressants have been shown to be more effective compared to placebo in treatment of MDD, with effect sizes ranging from 1.15-1.55^2^. Interestingly, combinations of antidepressants provide more benefit in MDD than monotherapy^3^. Regardless, there is an unmet need for treatment of major depressive disorder, given chances of relapse and treatment resistance^4,5^. A treatment option for treatment resistant MDD is neuromodulation which encompasses interventions such as electroconvulsive therapy (ECT) and transcranial magnetic stimulation (TMS). ECT has shown a pooled response rate of 60-80% and a pooled remission rate of 50-60% for MDD^6^. However, ECT imposes barriers encompassing lack of equitable access as well as illness characteristics needed to receive it^7^. In addition to these, ECT also carries risk of transient cognitive adverse effects^8^.

Although less effective than ECT, TMS does not have cognitive adverse effects and consequently greater patient preference^9-11^. TMS is generally safe and well-tolerated^12^. Previous studies with ECT and continuation pharmacotherapy have shown benefits of continuing psychotropics while receiving ECT in MDD^13-15^. There are no consensus guidelines on dosing strategies with antidepressants that patients take when considering TMS for MDD. Hence it becomes critical to think about strategies to combine TMS and antidepressants, such that benefits of both treatments are leveraged for patients.

The mechanisms of both pharmacotherapy and TMS in the treatment of psychiatric disorders are at least partially based on changes in neuroplasticity^16^. Although antidepressants are known to be modulators of neurotransmitters such as serotonin, dopamine, and noradrenaline^17^, modulation of neurotransmitters is not sufficient to completely explain their mechanism of action. For example, administration of antidepressants results in a rapidly increased concentration of serotonin whereas the peak clinical efficacy of antidepressants happens weeks to months after initial administration^18^. Other proposed explanations about the mechanism of action are changes in network connectivity and neuroplasticity. For example, antidepressants have been shown to have synaptogenic, neurogenic, and neuroprotective effects in their treatment of psychiatric disorders^16^. When used as an investigative tool, various single pulse TMS paradigms have demonstrated that antidepressants alter cortical excitability and neuronal long-term potentiation (LTP) and long-term depression (LTD)^19^.

TMS affects neuronal plasticity by modulating long and short-term potentiation of neurons^20^. Because both TMS and antidepressants affect neuroplasticity, perhaps their interactions in treating MDD may plausibly have an additive or synergistic effect. An example of this interaction between brain stimulation and pharmacotherapy has been already showcased by combining clozapine and ECT, which led to remission of symptoms in patients with treatment resistant schizophrenia, that was only partially responsive to clozapine^21^.

Two previous systematic reviews examined whether active TMS differentiated from sham TMS as an augmentation agent with antidepressant medications in decreasing HAMS scores in MDD^22,23^. These systematic reviews encompassed seven and nine RCTs respectively^22,23^. A review encompassing seven RCTs performed a meta-analysis and showed a standardized mean difference (SMD) of 0.86 when comparing decrease in HAMD scores between active TMS combined with antidepressants compared with sham TMS combined with antidepressants^22^. Active TMS was superior to sham TMS in decreasing HAMD scores, when both were combined with antidepressant medications. The other systematic review encompassing nine RCTs also found a superior benefit for active TMS compared to sham TMS, when combined with antidepressant medications^23^.

Although both reviews showed active TMS combined with antidepressant medications to be superior to sham TMS combined with antidepressant medications, RCTs included in both reviews were heterogenous in their TMS parameters and session numbers. In addition, these reviews included studies wherein patients in both arms had been on antidepressant medications before TMS was added on as augmentation. Therefore, to examine the effect of the combination on depressive symptoms in a systematic fashion, it becomes pertinent to examine RCTs wherein both treatment options were administered simultaneously and dosed uniformly to patients. A review of this literature can help optimize our understanding of an interaction, if present, and advocate for clinical translation of the combination. The primary objective of this review is to summarize evidence from studies that combined TMS and antidepressants in a rigorous manner, in treatment of MDD. As a secondary objective, we also examined studies that used TMS as an augmenting agent to antidepressant medications but were not included in previous systematic reviews that examined the same.

## Methods

PubMed/Medline and PsycINFO were searched using the terms “transcranial magnetic stimulation” or “theta burst stimulation” along with terms denoting psychiatric drug classes (e.g., “SSRI,” “antipsychotic,” etc.) as well as individual psychiatric drug names (e.g., “fluoxetine,” “clozapine,” etc.). Drug classes that have utility in treatment of MDD (antidepressants, antipsychotics, mood stabilizers and benzodiazepines) were included in our search terms. Search terms used are listed in the supplementary material. PRISMA guidelines were followed to guide inclusion criteria for the review^24^. All studies were published between January 1980 and September 1^st^, 2022. All studies were published in English and screened by title, abstract, and full text by two authors (PC and GR) before a decision was made to include or exclude the study.

For the systematic review, we included studies that met the following inclusion criteria: 1) Investigating TMS as an adjunctive or augmentation agent with antidepressant medications; or administered concurrently with antidepressant medication titration with a comparator placebo arm; 2) Participants in the study met criteria for major depressive disorder (MDD) as defined by any edition of the Diagnostic Statistical Manual of Disorders (DSM) or International Classification of Diseases (ICD); 3) The study used quantitative measures that assess symptom improvement in MDD (e.g., Hamilton Depression Rating Scale); 4) the study involved adult participants ≥ 18 years of age; and 5) the study needed to be an RCT, open label trial or a retrospective chart review. For the review, we excluded case reports and case series. We also excluded studies which administered either TMS or antidepressants as maintenance treatment for MDD in a non-concurrent fashion. For the meta-analysis, we excluded studies which did not administer TMS and antidepressants concurrently.

In the meta-analysis, the primary outcome measure studied was reduction of symptom severity as measured by the Hamilton Depression Rating Scale (HDRS) for MDD. A random effects model was used because high heterogeneity between studies was expected. Effect size was calculated for the RCTs as described by Morris^25^, who argued that the pooled pretest standard deviation be used for weighting the differences of pre-post-means. Meta-analyses and calculations of heterogeneity (I^2^) were performed using RevMan 5.4^26^. Publication bias was assessed using Egger’s test for funnel plot asymmetry^27^ (using ‘metabias’ from meta package in Rstudio), since we had 10 studies in the meta-analysis to provide sufficient power.

## Results

From 731 unique articles that were screened, 30 studies were included in the qualitative synthesis. Figure 1 shows the PRISMA diagram of how selected studies were included in the review and meta-analyses. Studies we excluded comprised:- 1) experimental human TMS studies measuring cortical excitability in response to psychotropic medications (n=87), 2) clinical trials combining TMS and antidepressants in conditions other than MDD [such as obsessive compulsive disorder (OCD), tinnitus, migraine] (n=7)], 3) previous systematic reviews on utility of TMS for various disorders (n=50), 4) TMS combined with psychotropics other than antidepressants, including antipsychotics and benzodiazepines (n=19) and 5) animal studies utilizing electroconvulsive stimulation (ECS) (n=100). We assessed full texts of 31 articles for inclusion and excluded two studies. One article was excluded from this, and was a TMS study that used mirtazapine for maintenance treatment after course of TMS^28^. The final list included 30 studies. Table 1 summarizes key characteristics of each of these studies. Egger’s test for publication bias was not significant indicating a lack of publication bias for studies included in the meta-analysis [t(8)=1.92, p = 0.09] (See Figure 2).

**Figure 1.**
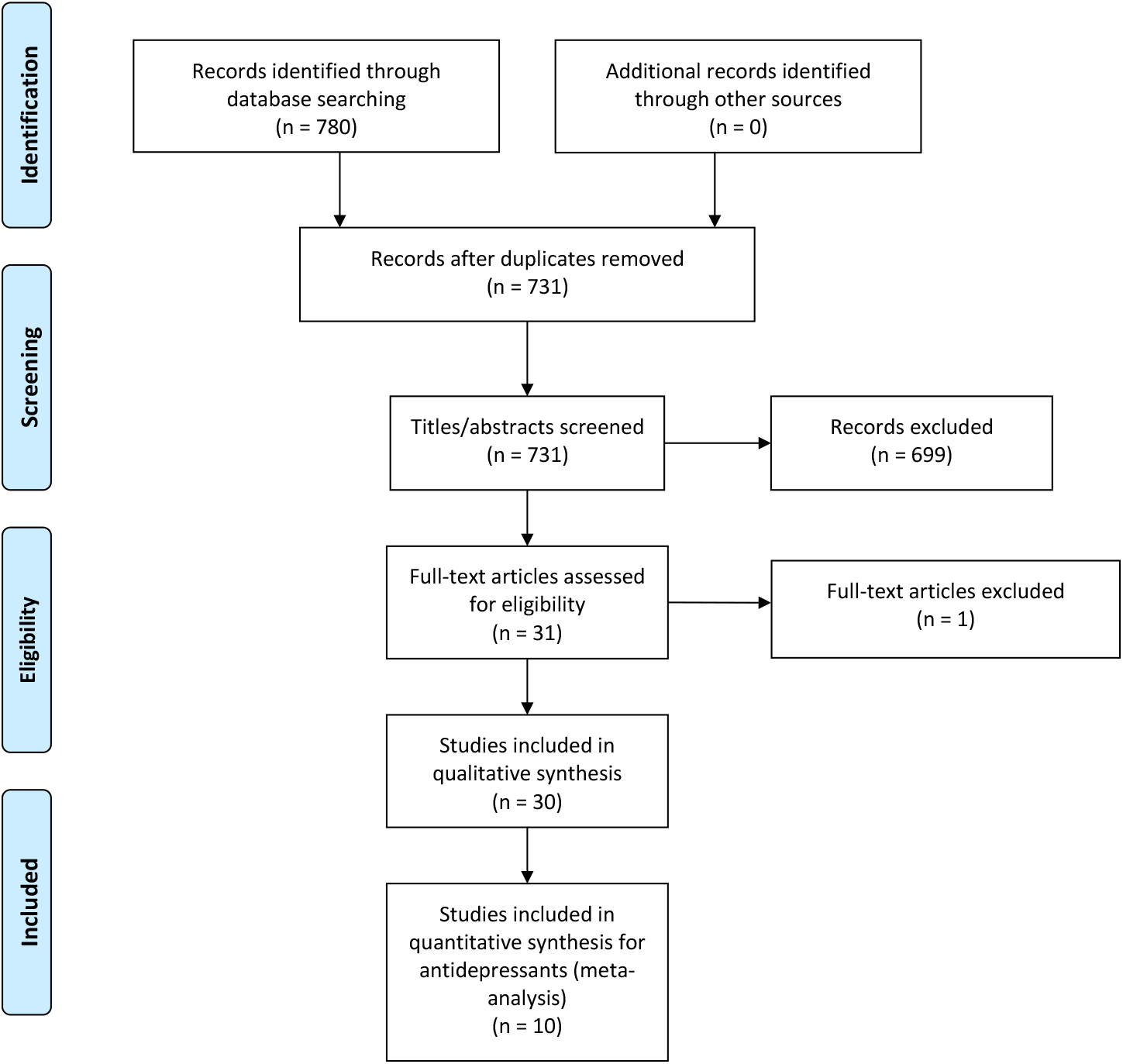
PRISMA diagram of the study selection process

**Figure 2.**
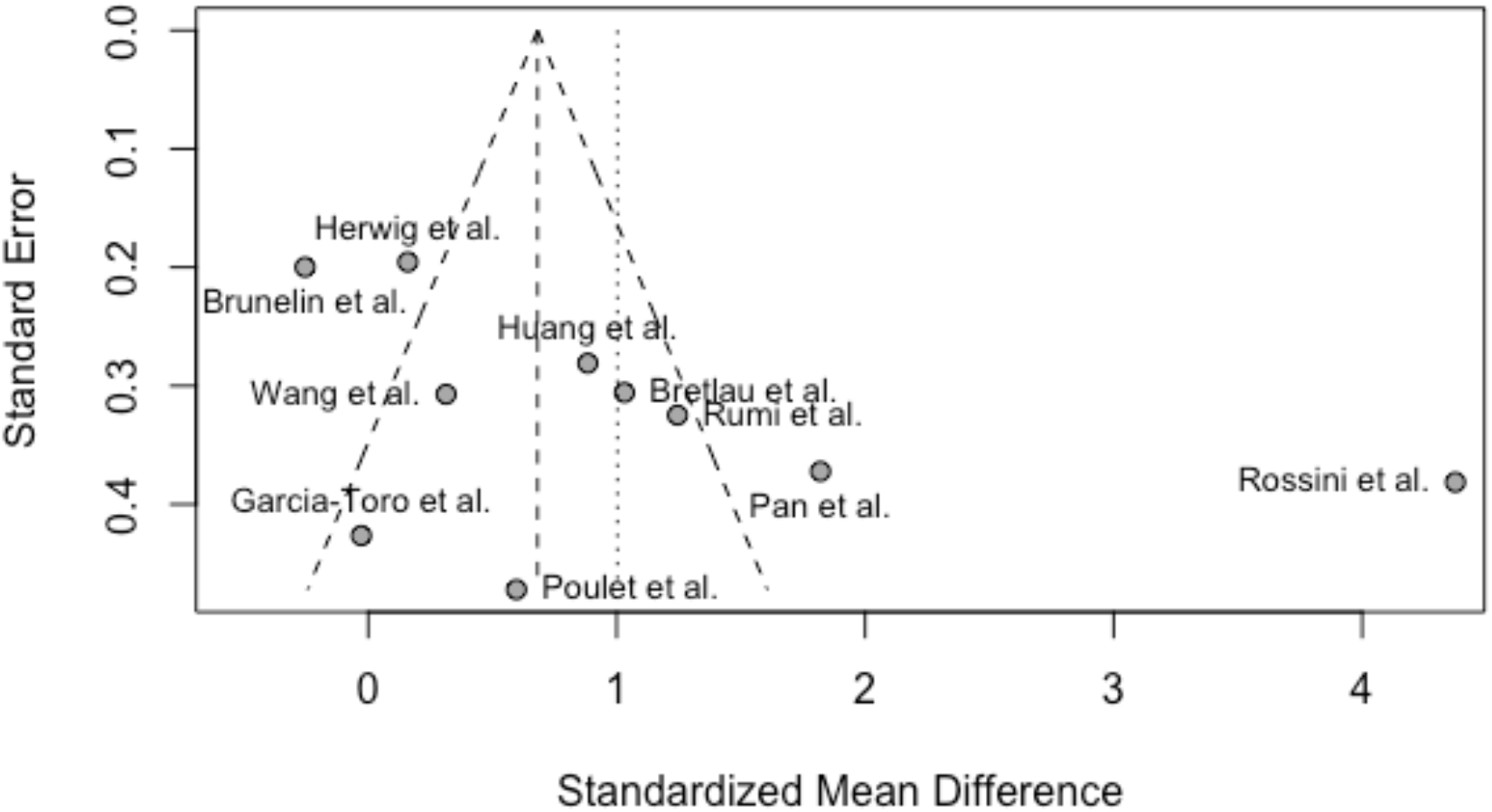
Funnel Plot of Standard Error by Standardized Mean Difference (Publication Bias)

**Table 1.**
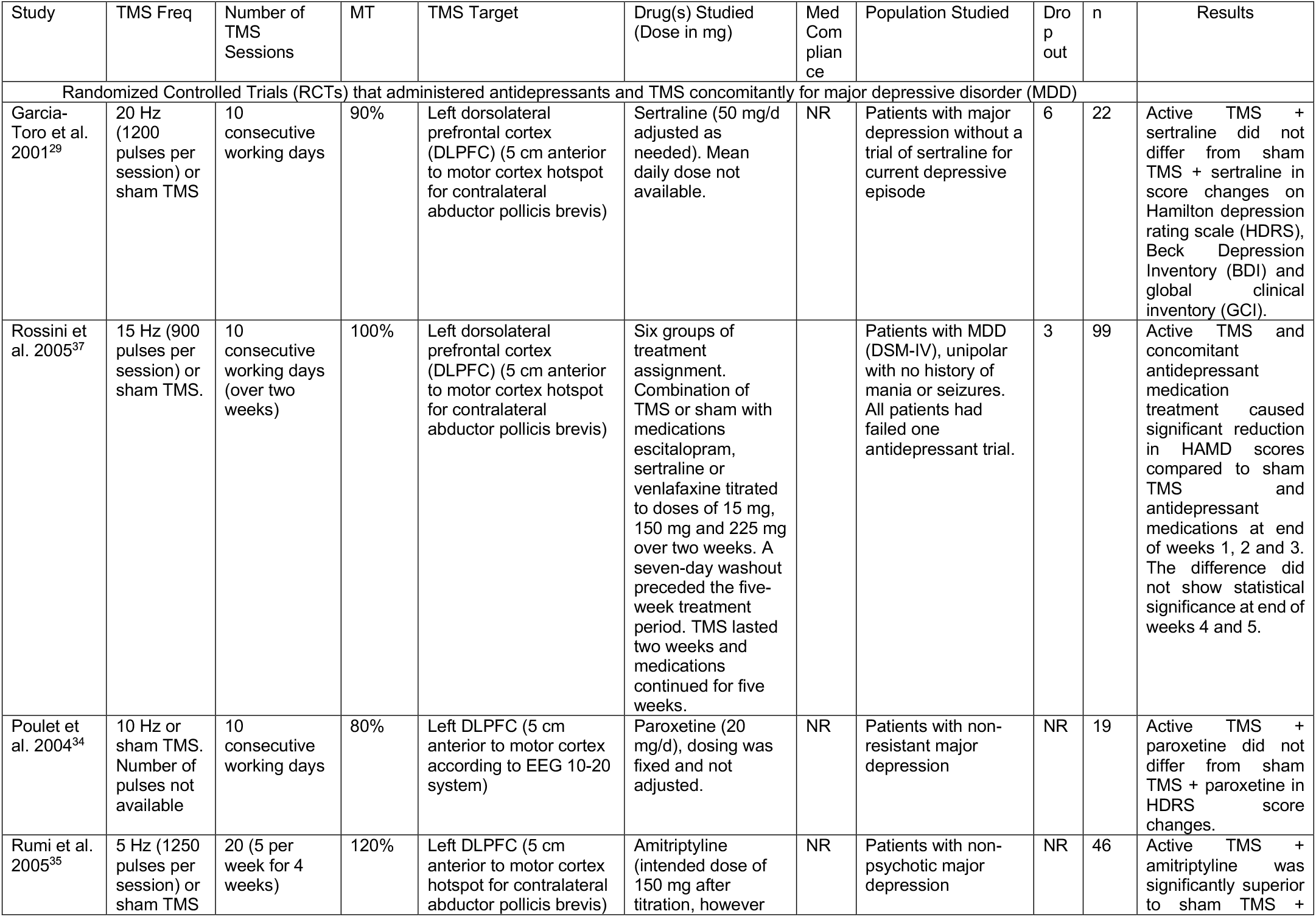
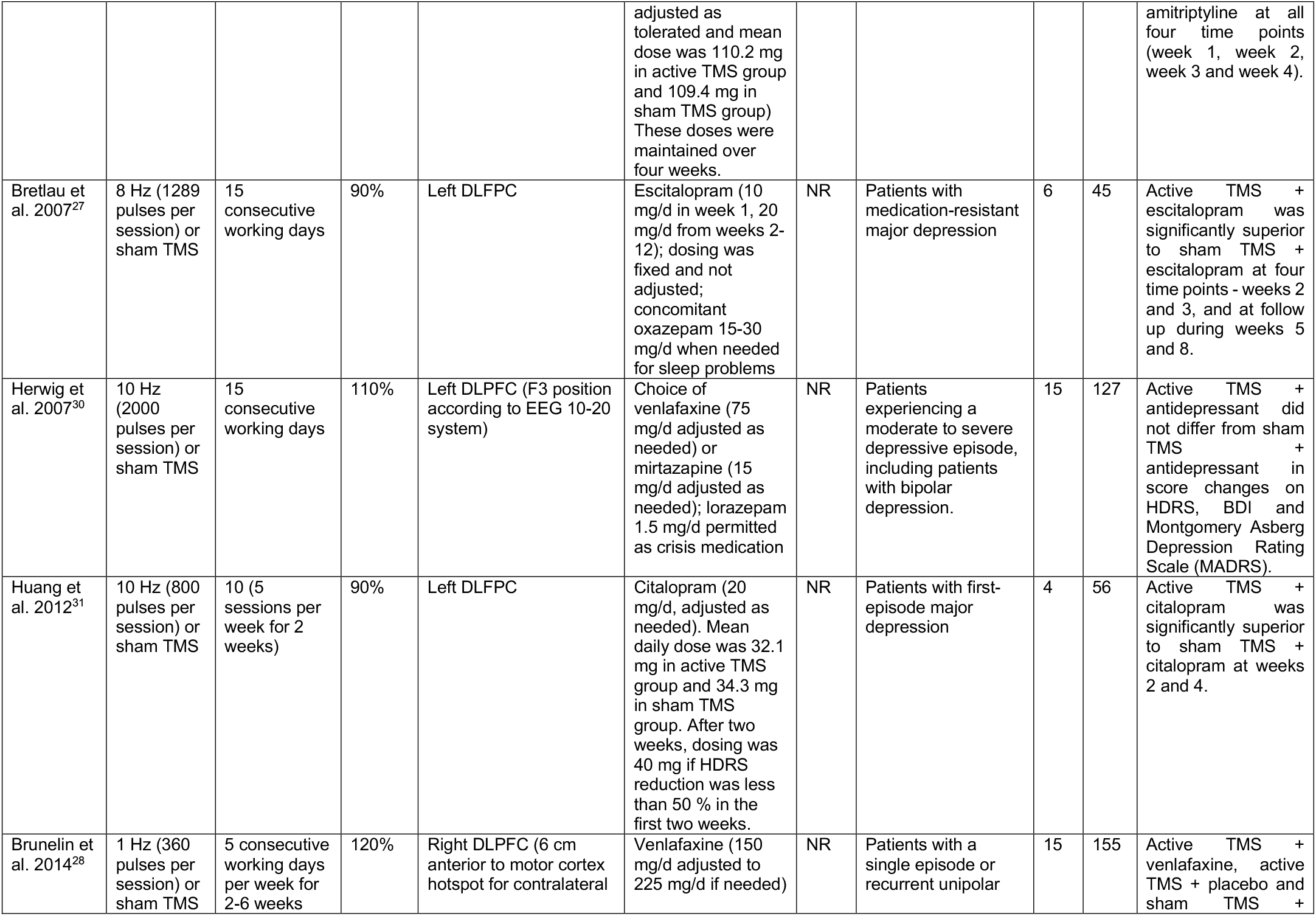
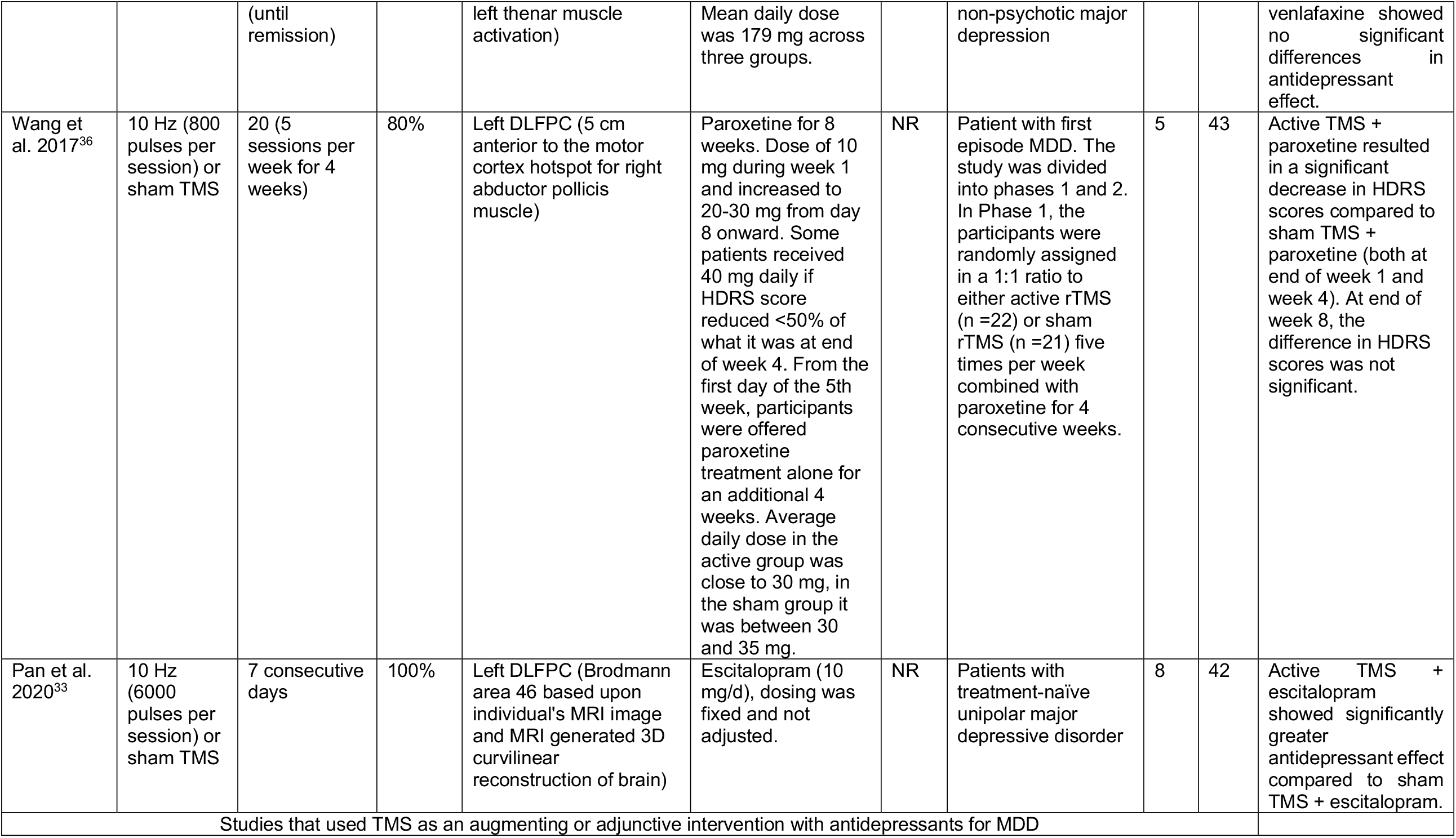
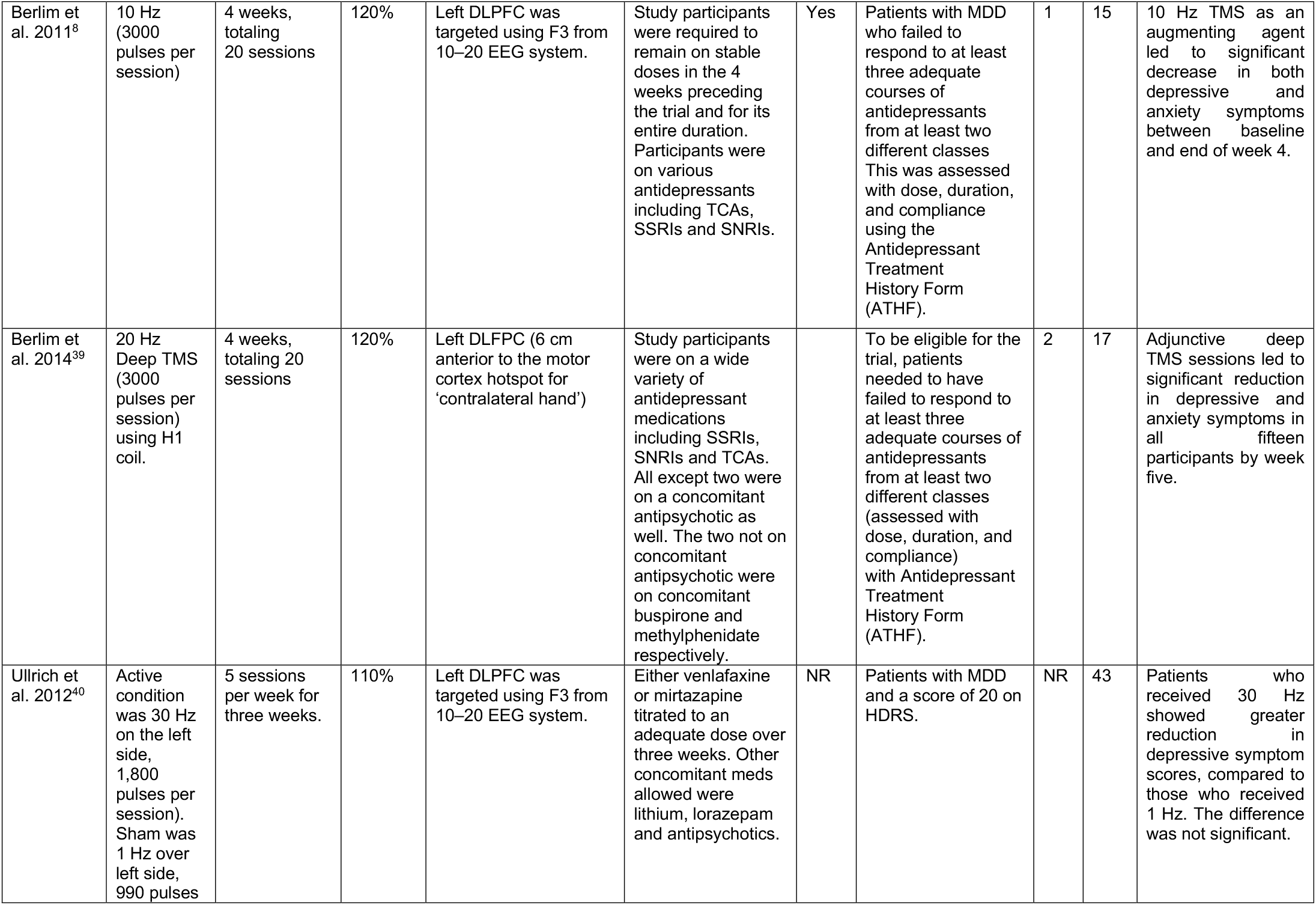
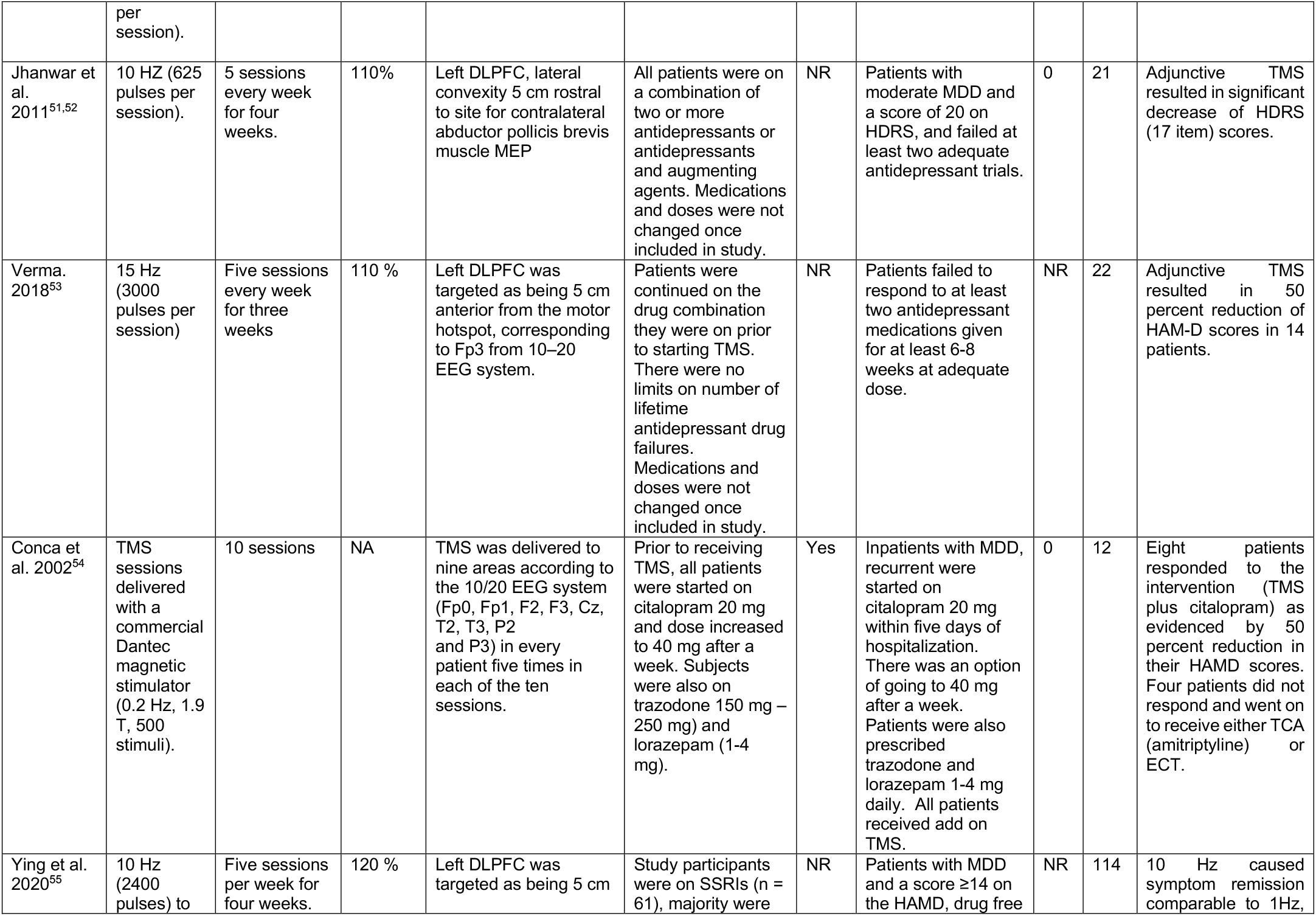
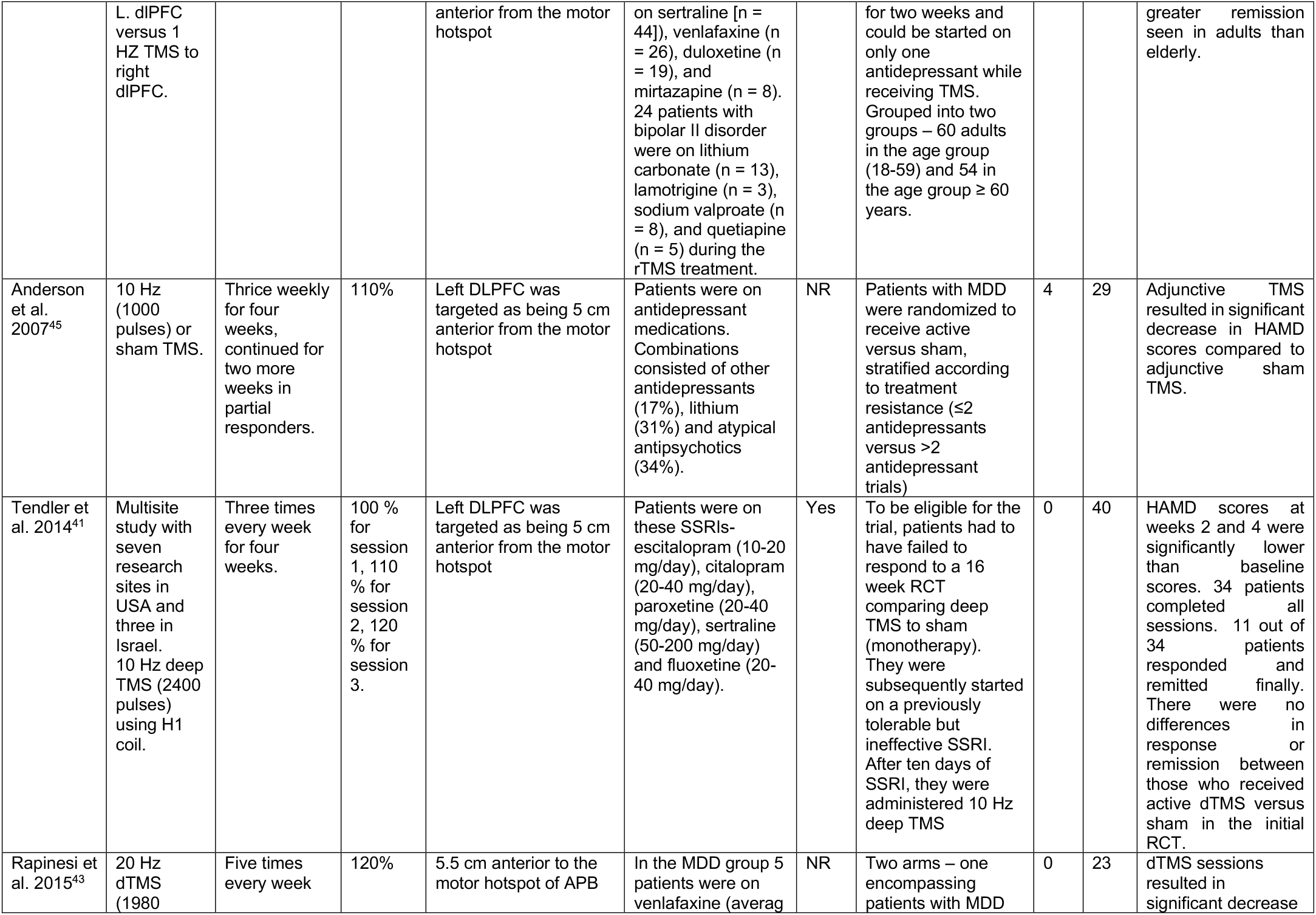
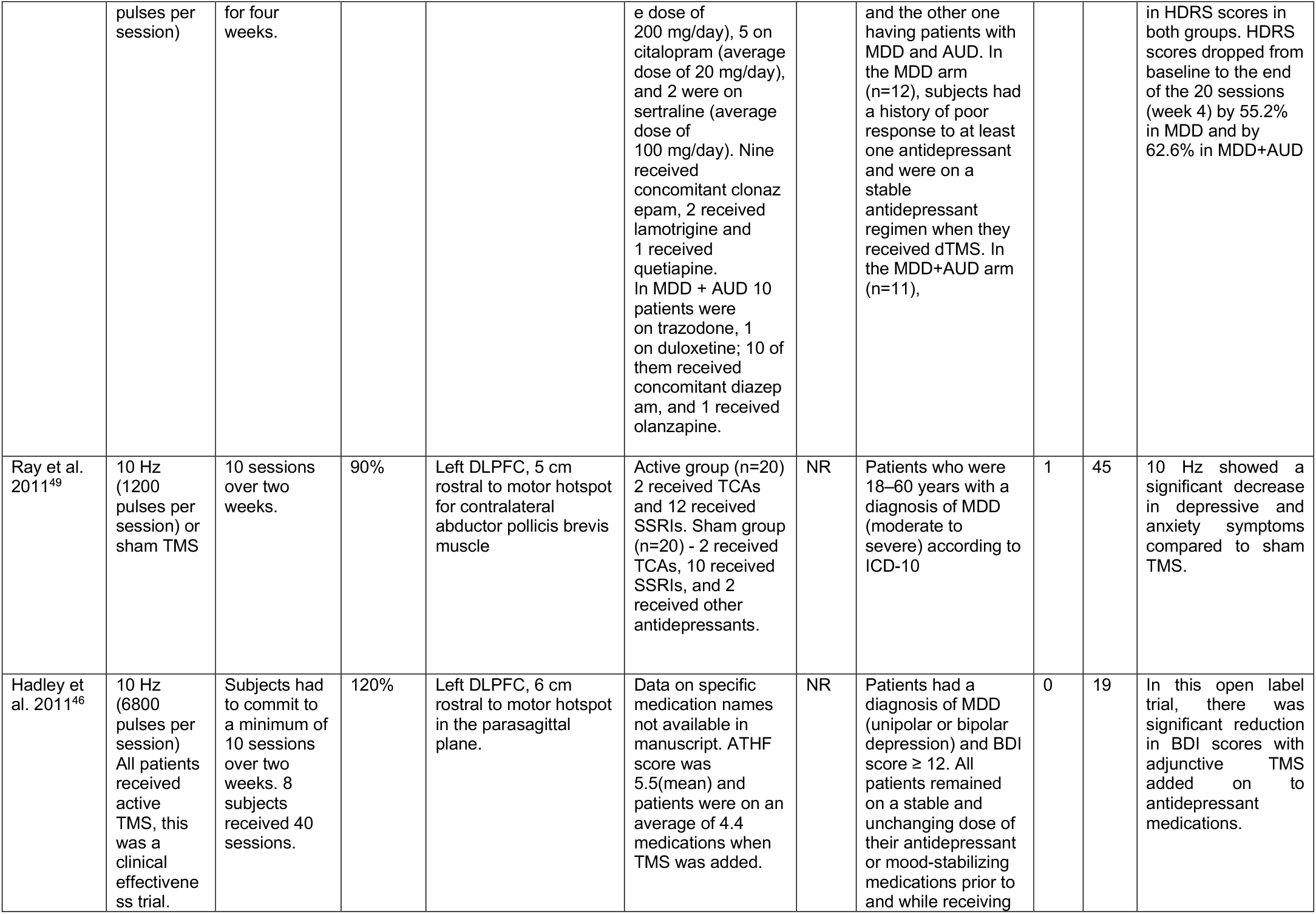
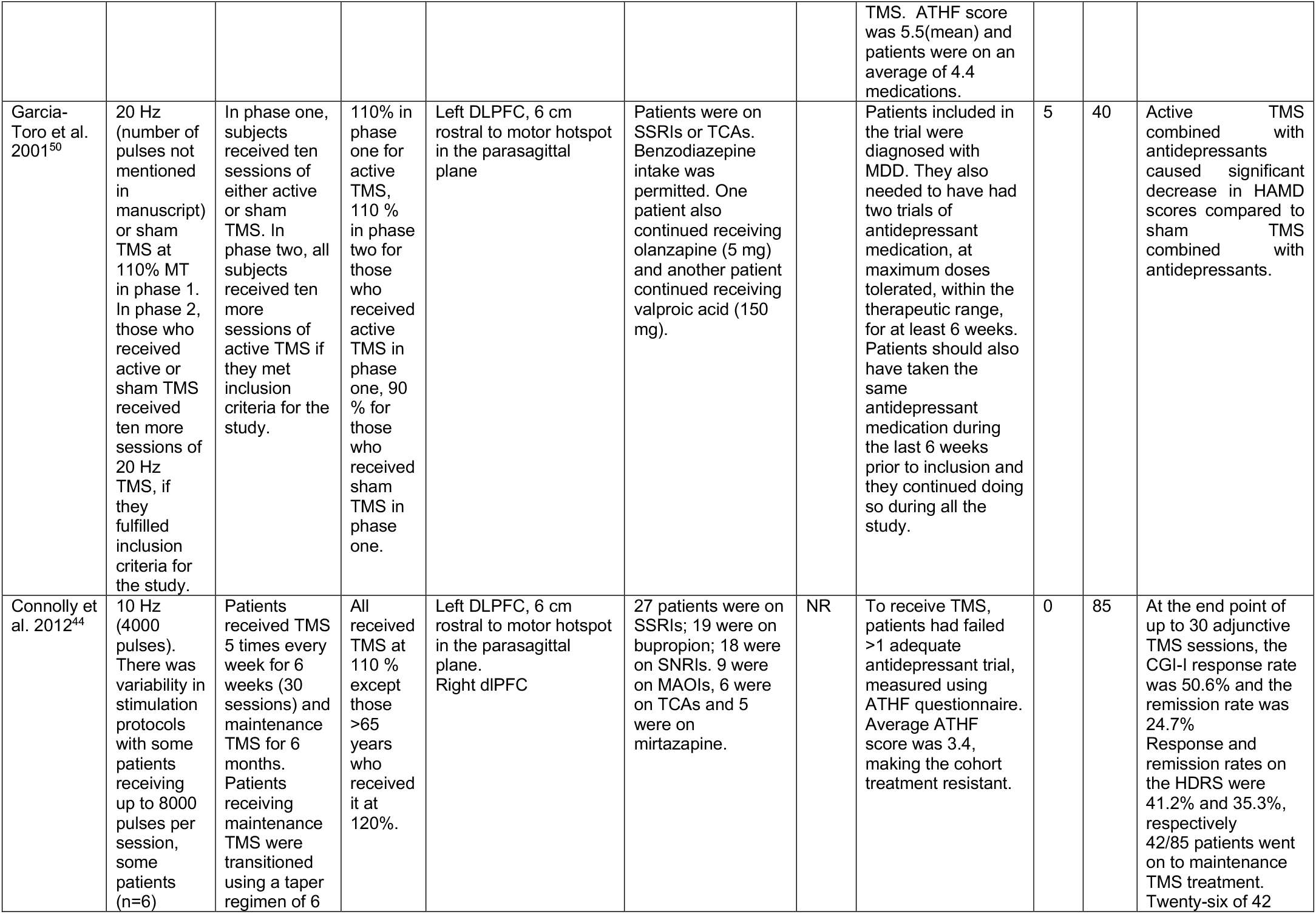
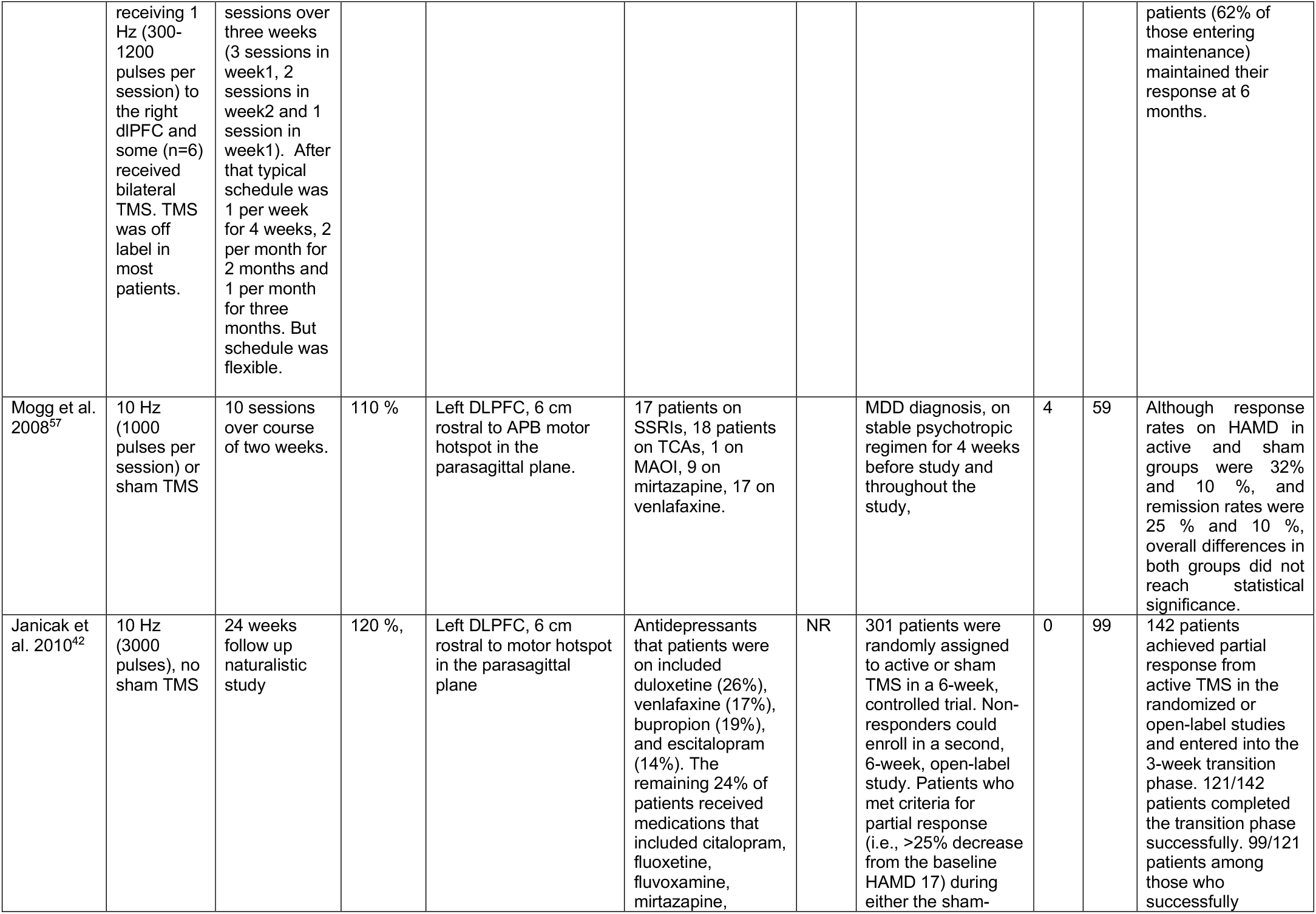
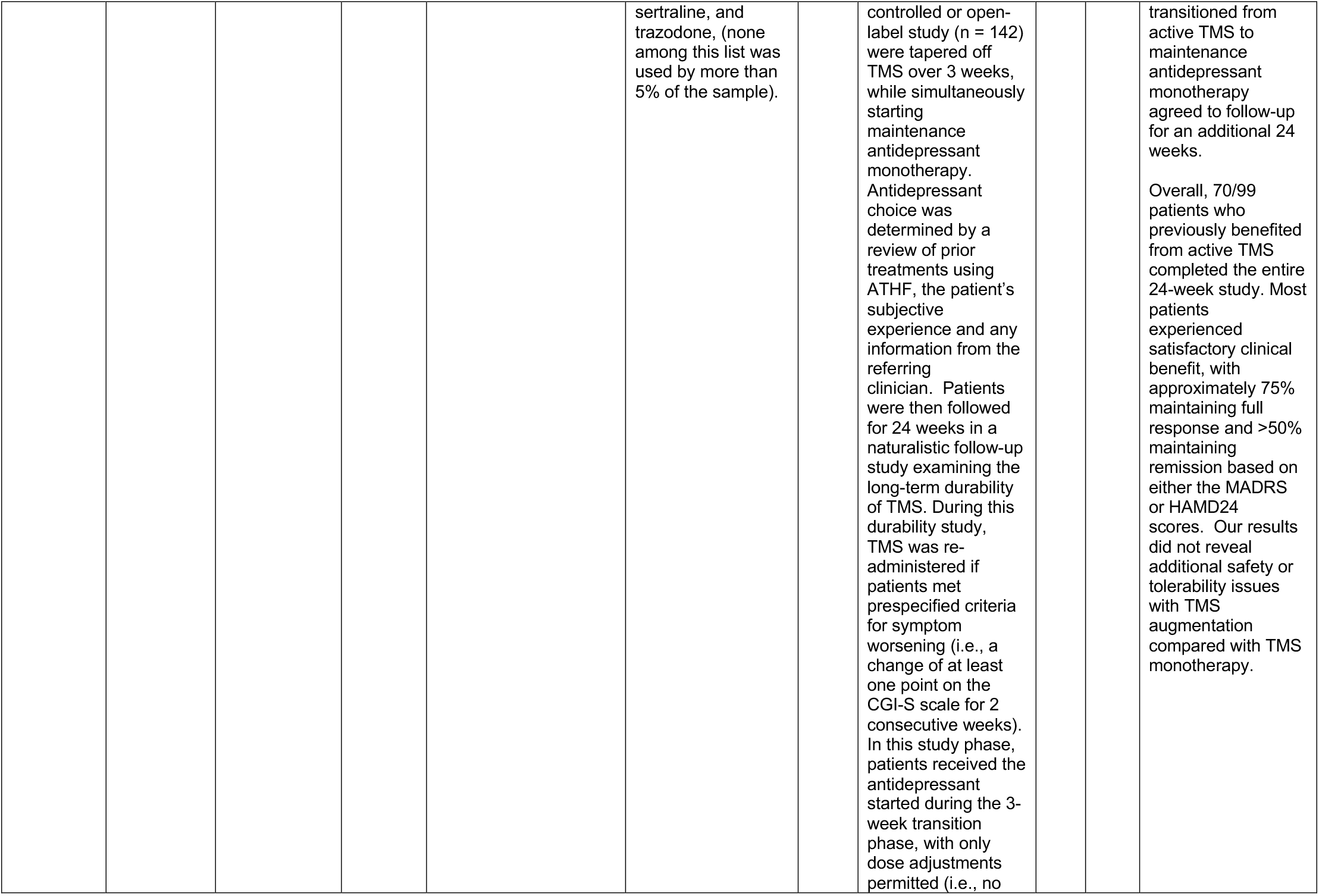
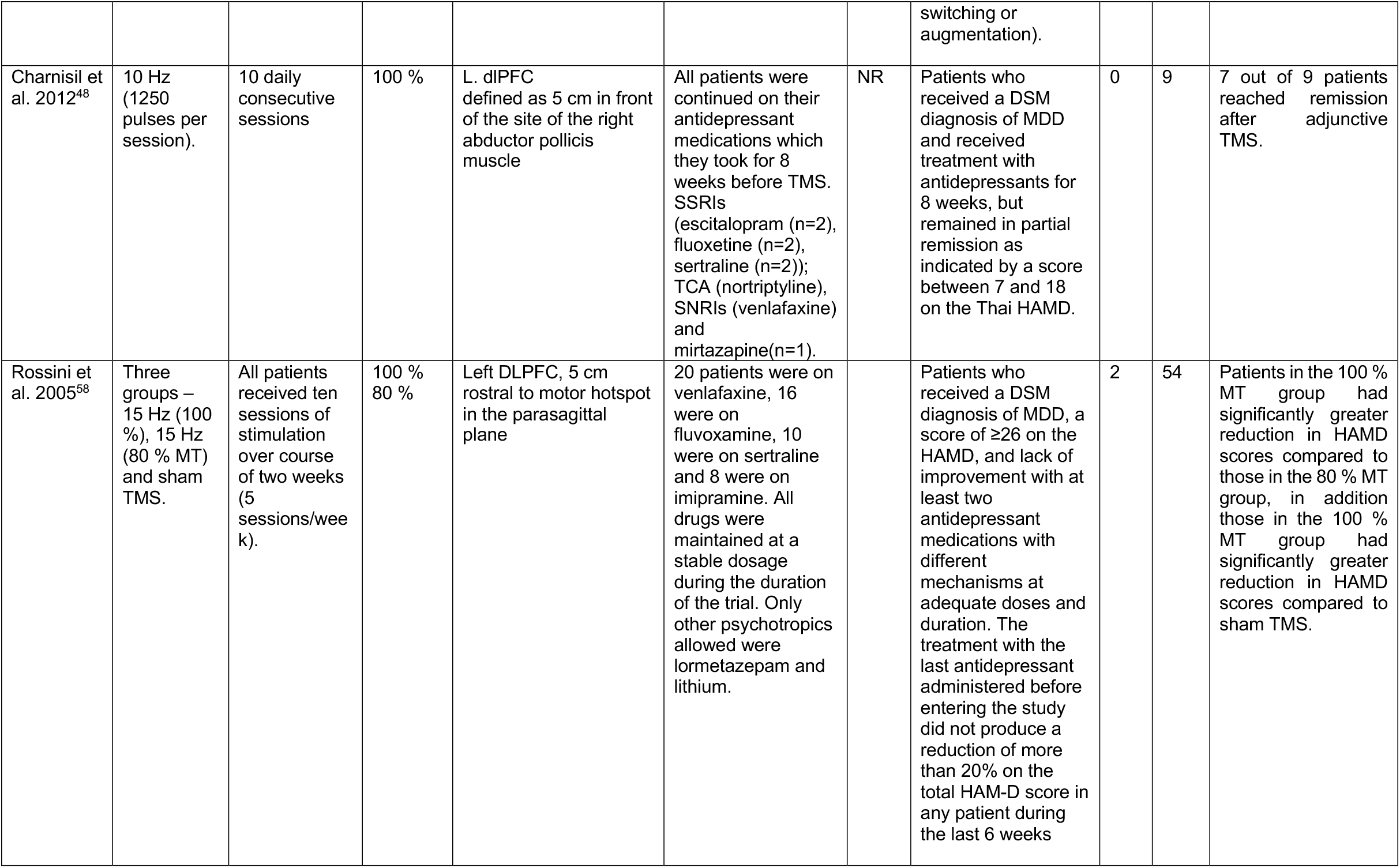
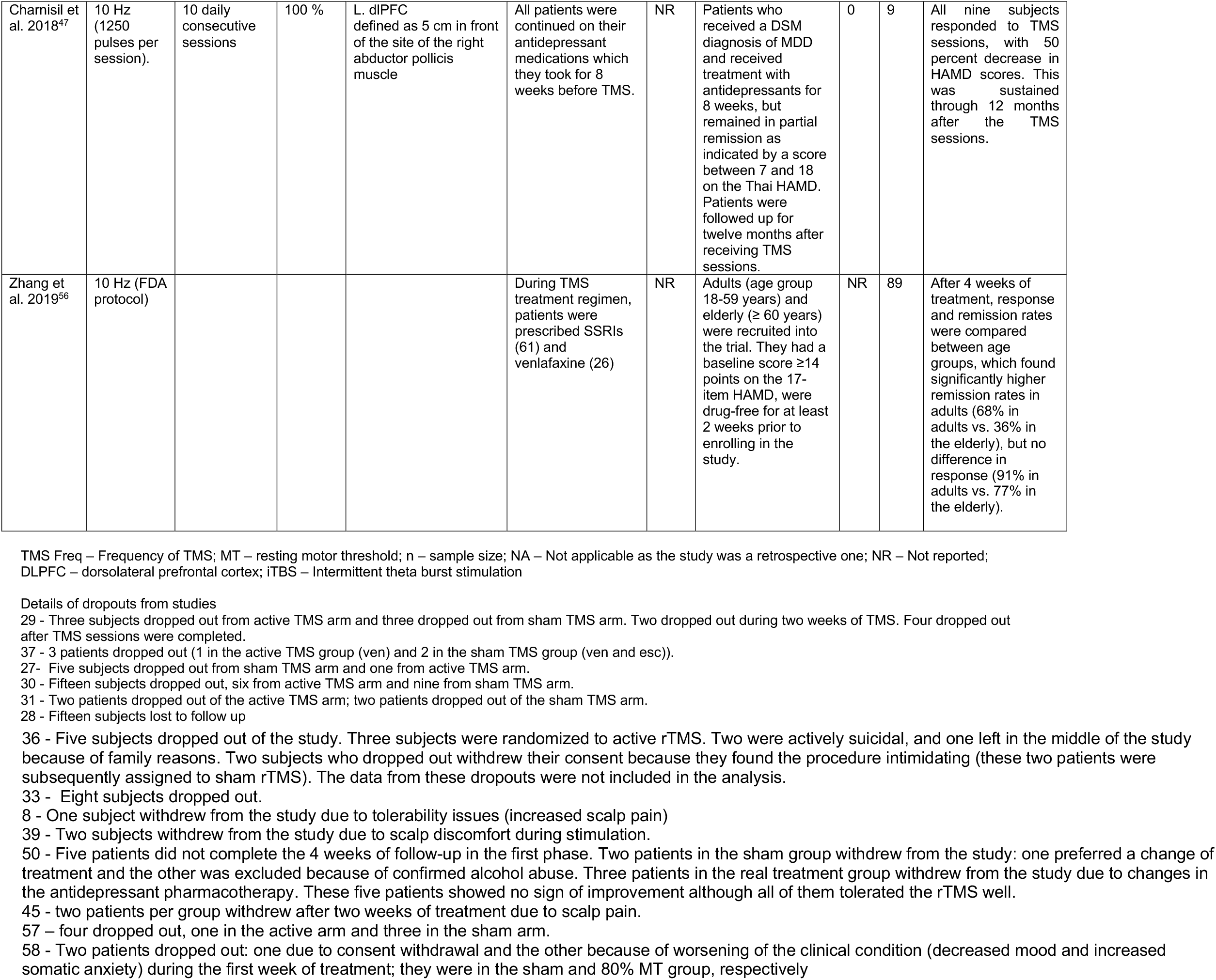
Summary of included studies

### TMS and Antidepressants

Ten randomized controlled trials (RCTs) were included in the meta-analysis. They investigated the combined use of TMS with antidepressants^29-38^. Specifically, six of them combined selective serotonin reuptake inhibitors (SSRIs) (encompassing citalopram, paroxetine, and escitalopram) with TMS^29,31,33,35,36,38^, two combined a serotonin-norepinephrine reuptake inhibitors (SNRI) (venlafaxine) with TMS^30,32^, one compared TMS with escitalopram to TMS with venlafaxine and TMS with sertraline^39^ and one combined amitriptyline with TMS^37^. Three RCTs used fixed doses of antidepressants with no adjustments during the study^29,35,36^.Scales used to measure outcomes included Hamilton Depression Rating Scale (HDRS), Montgomery-Asberg Depression Rating Scale (MADRS), and Beck Depression Inventory (BDI). Six out of ten RCTs reporting the antidepressant efficacy of the two groups found the combined use of antidepressants with active TMS resulted in significantly lower scores on depression scales and faster onset of antidepressant effect compared to antidepressants with sham TMS.

Two RCTs used low frequency TMS (1 Hz and 5 Hz)^30,37^ and all others used high frequency TMS (10-20 Hz)^29,31-33,35,36,38,39^. Number of sessions ranged from 5-30, with two studies delivering 5 and 7 sessions respectively^30,35^ and all others delivering 10 or more sessions^29,31-33,35-39^.

Ten RCTs investigated whether combined TMS with antidepressants had a greater efficacy for the treatment of depressive symptoms compared to combined sham TMS with antidepressants^29-33,35-39^. Thus, a meta-analysis was conducted of these RCTs. A common measure used between the studies was needed to conduct the meta-analysis. The change in HDRS was used as the outcome measure for the meta-analysis because it was the measure most used across studies. Poulet et al^36^ and Rumi et al^37^ did not report HDRS scores, but they did report MADRS scores. Therefore, to incorporate these studies into the meta-analysis, MADRS score means, and standard deviations were converted to equivalent HDRS score means and standard deviations using the chart provided by Leucht et al^40^. Because Poulet et al^36^ only reported changes in depressive symptoms as percent changes relative to baseline MADRS scores, all values used in the meta-analysis were converted to percent changes relative to baseline HDRS scores to allow for incorporation of Poulet et al^36^ into the meta-analysis. Figure 3 shows the results of the meta-analysis.

**Figure 3.**
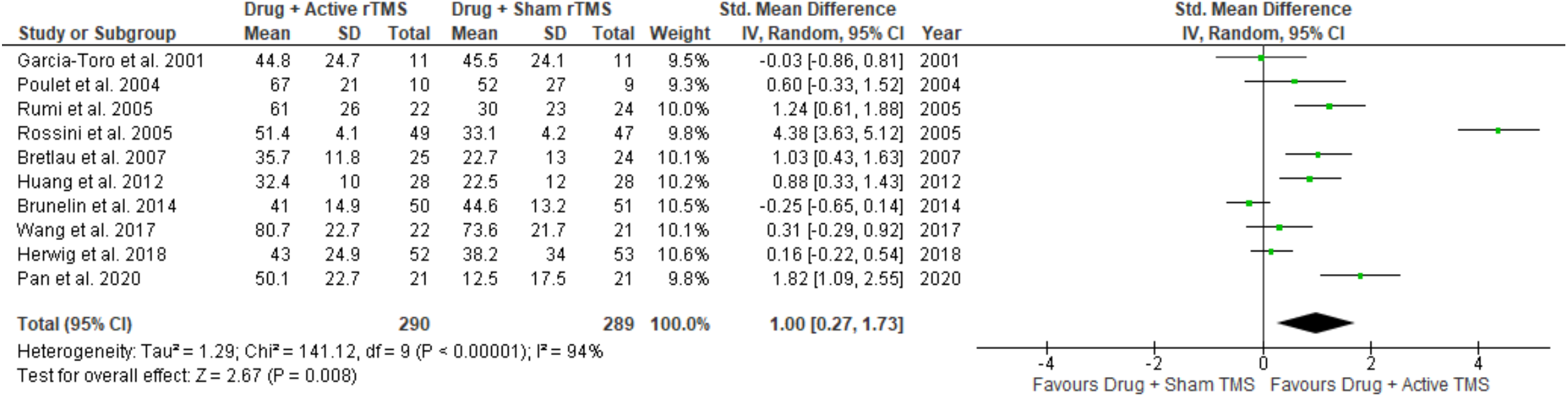
Meta-analysis of antidepressants combined with active versus sham TMS

The results of the random effects meta-analysis suggest antidepressants with active TMS had greater efficacy for the treatment of major depression compared to antidepressants with sham TMS (Hedge’s *g* = 1; 95% CI [0.27, 1.73]). As expected, there was a high heterogeneity between the studies (I^2^ = 94%; P < 0.00001). We assessed publication bias for included studies and did not find any [t (8) = 1.92, p=0.09).

### TMS as augmentation or adjunctive to antidepressant medications

We included 20 studies that administered TMS either as augmentation or as an adjunctive intervention to patients already on antidepressants^11,41-59^(Table 1). Of these twenty studies, seven were RCTs that compared active versus sham TMS as an adjunctive treatment modality, added on to antidepressant regimens that study participants were already on^42,47,51,52,57,59,60^. Three studies were clinical effectiveness trials that modeled real world scenarios combining TMS with antidepressant medications^44,46,48^. Only five studies used the antidepressant treatment history form (ATHF)^61^ to assess treatment resistance and ensured this as a for patients to participate in these trials^11,41,44,46,48^. All 20 studies showed significant reduction in depressive symptoms across patients. There was variability in TMS parameters, and except two studies^42,56^, none of the other trials had restrictions on medications that patients could be on to participate in the trial. Two studies restricted patients to take either venlafaxine or mirtazapine with a specific titration schedule^42^ or citalopram that was titrated in the same manner for all participants^56^. Nine out of twenty studies had patients who were on other psychotropic medications (mood stabilizers, antipsychotics and benzodiazepines) in addition to antidepressants.

Fewer patients in these trials dropped out or reported lack of tolerability to TMS in studies that used TMS as an adjunct intervention to antidepressants (n=19 across 13 studies), compared to RCTs that administered TMS and antidepressants concurrently (n=62 across 8 studies). All except two of these studies used L. dlPFC targeted as 5-6 cm in front of the motor hotspot^46,56^. None of these studies using neuroimaging for targeting. No study stratified patient outcomes based on antidepressants. Two trials that administered maintenance TMS if needed, showed sustained remission rates at 6 months after acute treatment^44,46^.

## Discussion

The current systematic review and meta-analysis reviewed the literature on 1) the efficacy of concurrently administered TMS with antidepressants for treatment of MDD and 2) effectiveness of augmenting antidepressants with TMS or using TMS as an adjunctive intervention to antidepressants in MDD.

Our meta-analysis found TMS combined with concurrent antidepressant therapy may have greater efficacy for the treatment of MDD compared to antidepressants combined with sham TMS. Since sham TMS uses a significantly weaker electric field compared to active TMS^62^, it would be pragmatic to state that the combination of TMS and antidepressants included in this review showed a large effect size compared to antidepressants alone. The findings of the meta-analysis, however, should be interpreted with a high degree of caution given the large amount of heterogeneity between the studies, encompassing experimental procedures (antidepressant class, TMS frequency, number of sessions, and stimulation intensity) used in the studies.

Clinical trials that augmented antidepressants with TMS or used TMS as an adjunctive intervention to antidepressants showed significant clinical benefit for patients who participated in the trials. Also notable is the fact that dropout rates were fewer in these trials compared to the RCTs included in the meta-analysis. Thirteen of these twenty studies were either naturalistic studies or open label studies without randomization or blinding. Naturalistic studies had greater flexibility in scheduling TMS sessions when participants missed sessions. This may have attributed to participant retention and decreased rate of drop out. These trials also modeled real world scenarios witnessed by clinicians treating treatment resistant MDD.

The review and meta-analysis have significant utility for clinicians, to validate the approach of using TMS as an augmenting agent or adjunctive intervention, while keeping patients on their antidepressants. Nonetheless, these studies also showcased some potential confounders such as variability in psychotropic regimens for patients with MDD who participated in these trials (having mood stabilizer medications and antipsychotics in addition to antidepressants), variability in definition of treatment resistant MDD and diagnostic variability (unipolar versus bipolar depression). We would like to emphasize that despite these confounders, patient retention was not a problem in these trials.

TBS has shown greater effect size compared to sham TMS (0.64) for treatment of MDD^63^. TBS is more efficient than other high frequency TMS protocols, in that it requires 3 to 9 minutes for administration compared to an hour for 10 Hz TMS. An accelerated intermittent TBS (iTBS) protocol called Stanford Neuromodulation Therapy (SNT) using ten daily sessions of intermittent TBS (iTBS) for five days has recently shown promise in the treatment of MDD as well^64,65^. In the first Stanford Neuromodulation Treatment (SNT) trial (n=20), participants were required to maintain their psychotropic medication regimen throughout the duration of the trial. Fourteen of the twenty participants in the trial were on antidepressant medications. Three of the remaining six were not on any psychotropic medications and the remaining three participants were on other medications^65^. This trial did not use the ATHF^65^. In the second SNT trial (n=29), 27 participants were on antidepressant medications (including SSRIs, SNRIs, MAOIs, TCAs and atypical ones including bupropion and mirtazapine). This trial used ATHF to track medication history of participants and the scores were similar between active and sham groups^64^.

An alternative strategy to increase effectiveness of iTBS could also be optimally combining iTBS with antidepressants for increasing response and remission rates in these illnesses. These prospective trials must initially collect history regarding previous psychotropic trials, similar to the ATHF for previous antidepressant trials^66^. They must also track compliance and seek to move towards consensus guidelines for dosing antidepressants and other psychotropics with iTBS.

There are already multiple examples in medicine where the use of combined treatment methods results in greater efficacy. For example, multiple antidiabetic drugs with different mechanisms are often administered simultaneously to elicit a greater drop in hemoglobin A1c than drug monotherapy^21,67^. In a seminal crossover trial, combining ECT and clozapine resulted in an initial 50% response in patients with treatment resistant schizophrenia, who received the combination and a 47% response rate in patients randomized to receive only clozapine initially but were then crossed over into the clozapine-ECT arm^21^. Support for the combination arose from independent and distinct effects exhibited by both treatment modalities on symptoms of schizophrenia.

Two studies that used maintenance TMS showed sustained remission rates. A previous study that combined antidepressants with maintenance TMS showed similar proportions of response, remission and partial response across patients on various classes of antidepressants (SSRI versus SNRIs versus TCAs versus MAOIs)^68^. Regardless of TMS parameters or number of sessions delivered acutely until response or remission is attained, relapse rates over time are high in MDD without maintenance TMS sessions^69,70^. Like the symptom titrated algorithm based longitudinal ECT (STABLE) regimen in ECT, it is essential to move towards consensus regimens for maintenance TMS protocols to sustain remission in MDD^12,71,72^. In the context of combining antidepressants and TMS, it is important to discuss maintenance TMS given various clinical scenarios that could arise when providing TMS for patients with MDD. These include maintenance treatment with antidepressants after the index course of TMS, combining maintenance treatments with TMS and antidepressants or even maintenance treatment with TMS while tapering antidepressants. Future research needs to include both parametric studies and RCTs that model interactions between antidepressants and TMS (both index courses and plausible maintenance regimens).

## Conclusion

There is limited evidence which suggests TMS combined with pharmacotherapy can have greater efficacy for the treatment of MDD. However, the conclusions which can be made from these findings are limited by the small number of studies included in the review, the small sample sizes within most of the studies, and heterogeneity between the studies. The limited evidence supports an increased benefit of combining TMS and antidepressants in MDD without increasing risk of adverse events.

Given the design of the trials we could choose for our meta-analysis, it is not essentially clear if the effect we saw is due to use of active versus sham TMS or an interaction between TMS and medication. In order to tease this out, the ideal strategy would be comparing a combination of active TMS and a select antidepressant therapy with a combination of active TMS and placebo. Both parametric studies to study the interaction and RCTs comparing active TMS plus antidepressant versus active TMS plus placebo are needed to systematically examine how to combine TMS and antidepressants. These trials can use the ATHF to assess treatment resistance and use currently available naturalistic studies to model dosing and regimens.

## Data Availability

Systematic review/meta-analysis

## Acknowledgment

This work was supported by the National Institutes of Health grant numbers AA026255 (CRR), TR001997 (CRR), CA225419 (SSH) and University of Kentucky College of Medicine (GR). We would like to thank Jackson L. Weber for help with figures.

## References

1. Friedrich MJ. Depression Is the Leading Cause of Disability Around the World. JAMA. 2017; 317(15):1517.

2. Cipriani A, Furukawa TA, Salanti G, et al. Comparative efficacy and acceptability of 21 antidepressant drugs for the acute treatment of adults with major depressive disorder: a systematic review and network meta-analysis. Lancet. 2018; 391(10128):1357–1366.

3. Henssler J, Alexander D, Schwarzer G, et al. Combining Antidepressants vs Antidepressant Monotherapy for Treatment of Patients With Acute Depression: A Systematic Review and Meta-analysis. JAMA Psychiatry. 2022; 79(4):300–312.

4. Hasin DS, Goodwin RD, Stinson FS, et al. Epidemiology of major depressive disorder: results from the National Epidemiologic Survey on Alcoholism and Related Conditions. Arch Gen Psychiatry. 2005; 62(10):1097–1106.

5. Kessler RC, Berglund P, Demler O, et al. The epidemiology of major depressive disorder: results from the National Comorbidity Survey Replication (NCS-R). JAMA. 2003; 289(23):3095–3105.

6. Espinoza RT, Kellner CH. Electroconvulsive Therapy. N Engl J Med. 2022; 386(7):667–672.

7. Wilkinson ST, Kitay BM, Harper A, et al. Barriers to the Implementation of Electroconvulsive Therapy (ECT): Results From a Nationwide Survey of ECT Practitioners. Psychiatr Serv. 2021; 72(7):752–757.

8. Porter RJ, Baune BT, Morris G, et al. Cognitive side-effects of electroconvulsive therapy: what are they, how to monitor them and what to tell patients. BJPsych Open. 2020; 6(3):e40.

9. Magnezi R, Aminov E, Shmuel D, et al. Comparison between neurostimulation techniques repetitive transcranial magnetic stimulation vs electroconvulsive therapy for the treatment of resistant depression: patient preference and cost-effectiveness. Patient Prefer Adherence. 2016; 10:1481–1487.

10. Berlim MT, Van den Eynde F, Daskalakis ZJ. Efficacy and acceptability of high frequency repetitive transcranial magnetic stimulation (rTMS) versus electroconvulsive therapy (ECT) for major depression: a systematic review and meta-analysis of randomized trials. Depress Anxiety. 2013; 30(7):614–623.

11. Berlim MT, McGirr A, Beaulieu MM, et al. High frequency repetitive transcranial magnetic stimulation as an augmenting strategy in severe treatment-resistant major depression: a prospective 4-week naturalistic trial. J Affect Disord. 2011; 130(1-2):312–317.

12. Perera T, George MS, Grammer G, et al. The Clinical TMS Society Consensus Review and Treatment Recommendations for TMS Therapy for Major Depressive Disorder. Brain Stimul. 2016; 9(3):336–346.

13. Sackeim HA, Haskett RF, Mulsant BH, et al. Continuation pharmacotherapy in the prevention of relapse following electroconvulsive therapy: a randomized controlled trial. JAMA. 2001; 285(10):1299–1307.

14. Kellner CH, Husain MM, Knapp RG, et al. Right Unilateral Ultrabrief Pulse ECT in Geriatric Depression: Phase 1 of the PRIDE Study. Am J Psychiatry. 2016; 173(11):1101–1109.

15. Lisanby SH, McClintock SM, McCall WV, et al. Longitudinal Neurocognitive Effects of Combined Electroconvulsive Therapy (ECT) and Pharmacotherapy in Major Depressive Disorder in Older Adults: Phase 2 of the PRIDE Study. Am J Geriatr Psychiatry. 2022; 30(1):15–28.

16. Paulzen M, Veselinovic T, Grunder G. Effects of psychotropic drugs on brain plasticity in humans. Restor Neurol Neurosci. 2014; 32(1):163–181.

17. Stahl S. Stahl’s essential psychopharmacology: neuroscientific basis and practical application. 4th ed. Cambridge: Cambridge University Press; 2013.

18. Liu B, Liu J, Wang M, et al. From Serotonin to Neuroplasticity: Evolvement of Theories for Major Depressive Disorder. Front Cell Neurosci. 2017; 11:305.

19. Minzenberg MJ, Leuchter AF. The effect of psychotropic drugs on cortical excitability and plasticity measured with transcranial magnetic stimulation: Implications for psychiatric treatment. J Affect Disord. 2019; 253:126–140.

20. Klomjai W, Katz R, Lackmy-Vallee A. Basic principles of transcranial magnetic stimulation (TMS) and repetitive TMS (rTMS). Ann Phys Rehabil Med. 2015; 58(4):208–213.

21. Petrides G, Malur C, Braga RJ, et al. Electroconvulsive therapy augmentation in clozapine-resistant schizophrenia: a prospective, randomized study. Am J Psychiatry. 2015; 172(1):52–58.

22. Liu B, Zhang Y, Zhang L, et al. Repetitive transcranial magnetic stimulation as an augmentative strategy for treatment-resistant depression, a meta-analysis of randomized, double-blind and sham-controlled study. BMC Psychiatry. 2014; 14:342.

23. Brunoni AR, Fraguas R, Fregni F. Pharmacological and combined interventions for the acute depressive episode: focus on efficacy and tolerability. Ther Clin Risk Manag. 2009; 5:897–910.

24. Moher D, Liberati A, Tetzlaff J, et al. Preferred reporting items for systematic reviews and meta-analyses: the PRISMA statement. PLoS Med. 2009; 6(7):e1000097.

25. Morris SB. Estimating effect sizes from pretest-posttest-control group designs. Organizational Research Methods. 2008; 11(2):364–386.

26. Review Manager (RevMan). The Cochrane Collaboration; 2020.

27. Egger M, Davey Smith G, Schneider M, et al. Bias in meta-analysis detected by a simple, graphical test. BMJ. 1997; 315(7109):629–634.

28. Schule C, Zwanzger P, Baghai T, et al. Effects of antidepressant pharmacotherapy after repetitive transcranial magnetic stimulation in major depression: an open follow-up study. J Psychiatr Res. 2003; 37(2):145–153.

29. Bretlau LG, Lunde M, Lindberg L, et al. Repetitive transcranial magnetic stimulation (rTMS) in combination with escitalopram in patients with treatment-resistant major depression: a double-blind, randomised, sham-controlled trial. Pharmacopsychiatry. 2008; 41(2):41–47.

30. Brunelin J, Jalenques I, Trojak B, et al. The efficacy and safety of low frequency repetitive transcranial magnetic stimulation for treatment-resistant depression: the results from a large multicenter French RCT. Brain Stimul. 2014; 7(6):855–863.

31. Garcia-Toro M, Pascual-Leone A, Romera M, et al. Prefrontal repetitive transcranial magnetic stimulation as add on treatment in depression. J Neurol Neurosurg Psychiatry. 2001; 71(4):546–548.

32. Herwig U, Fallgatter AJ, Hoppner J, et al. Antidepressant effects of augmentative transcranial magnetic stimulation: randomised multicentre trial. Br J Psychiatry. 2007; 191:441–448.

33. Huang ML, Luo BY, Hu JB, et al. Repetitive transcranial magnetic stimulation in combination with citalopram in young patients with first-episode major depressive disorder: a double-blind, randomized, sham-controlled trial. Aust N Z J Psychiatry. 2012; 46(3):257–264.

34. Hunter AM, Minzenberg MJ, Cook IA, et al. Concurrent medication use and clinical outcome of repetitive Transcranial Magnetic Stimulation (rTMS) treatment of Major Depressive Disorder. Brain Behav. 2019; 9(5):e01275.

35. Pan F, Shen Z, Jiao J, et al. Neuronavigation-Guided rTMS for the Treatment of Depressive Patients With Suicidal Ideation: A Double-Blind, Randomized, Sham-Controlled Trial. Clin Pharmacol Ther. 2020; 108(4):826–832.

36. Poulet E, Brunelin J, Boeuve C, et al. Repetitive transcranial magnetic stimulation does not potentiate antidepressant treatment. Eur Psychiatry. 2004; 19(6):382–383.

37. Rumi DO, Gattaz WF, Rigonatti SP, et al. Transcranial magnetic stimulation accelerates the antidepressant effect of amitriptyline in severe depression: a double-blind placebo-controlled study. Biol Psychiatry. 2005; 57(2):162–166.

38. Wang YM, Li N, Yang LL, et al. Randomized controlled trial of repetitive transcranial magnetic stimulation combined with paroxetine for the treatment of patients with first-episode major depressive disorder. Psychiatry Res. 2017; 254:18–23.

39. Rossini D, Magri L, Lucca A, et al. Does rTMS hasten the response to escitalopram, sertraline, or venlafaxine in patients with major depressive disorder? A double-blind, randomized, sham-controlled trial. J Clin Psychiatry. 2005; 66(12):1569–1575.

40. Leucht S, Fennema H, Engel RR, et al. Translating the HAM-D into the MADRS and vice versa with equipercentile linking. J Affect Disord. 2018; 226:326–331.

41. Berlim MT, Van den Eynde F, Tovar-Perdomo S, et al. Augmenting antidepressants with deep transcranial magnetic stimulation (DTMS) in treatment-resistant major depression. World J Biol Psychiatry. 2014; 15(7):570–578.

42. Ullrich H, Kranaster L, Sigges E, et al. Ultra-high-frequency left prefrontal transcranial magnetic stimulation as augmentation in severely ill patients with depression: a naturalistic sham-controlled, double-blind, randomized trial. Neuropsychobiology. 2012; 66(3):141–148.

43. Tendler A, Gersner R, Roth Y, et al. Alternate day dTMS combined with SSRIs for chronic treatment resistant depression: A prospective multicenter study. J Affect Disord. 2018; 240:130–136.

44. Janicak PG, Nahas Z, Lisanby SH, et al. Durability of clinical benefit with transcranial magnetic stimulation (TMS) in the treatment of pharmacoresistant major depression: assessment of relapse during a 6-month, multisite, open-label study. Brain Stimul. 2010; 3(4):187–199.

45. Rapinesi C, Curto M, Kotzalidis GD, et al. Antidepressant effectiveness of deep Transcranial Magnetic Stimulation (dTMS) in patients with Major Depressive Disorder (MDD) with or without Alcohol Use Disorders (AUDs): a 6-month, open label, follow-up study. J Affect Disord. 2015; 174:57–63.

46. Connolly KR, Helmer A, Cristancho MA, et al. Effectiveness of transcranial magnetic stimulation in clinical practice post-FDA approval in the United States: results observed with the first 100 consecutive cases of depression at an academic medical center. J Clin Psychiatry. 2012; 73(4):e567–573.

47. Anderson IM, Delvai NA, Ashim B, et al. Adjunctive fast repetitive transcranial magnetic stimulation in depression. Br J Psychiatry. 2007; 190:533–534.

48. Hadley D, Anderson BS, Borckardt JJ, et al. Safety, tolerability, and effectiveness of high doses of adjunctive daily left prefrontal repetitive transcranial magnetic stimulation for treatment-resistant depression in a clinical setting. J ECT. 2011; 27(1):18–25.

49. Charnsil C, Suttajit S, Boonyanaruthee V, et al. Twelve-month, prospective, open-label study of repetitive transcranial magnetic stimulation for major depressive disorder in partial remission. Neuropsychiatr Dis Treat. 2012; 8:393–397.

50. Charnsil C, Suttajit S, Boonyanaruthee V, et al. An open-label study of adjunctive repetitive transcranial magnetic stimulation (rTMS) for partial remission in major depressive disorder. Int J Psychiatry Clin Pract. 2012; 16(2):98–102.

51. Ray S, Nizamie SH, Akhtar S, et al. Efficacy of adjunctive high frequency repetitive transcranial magnetic stimulation of left prefrontal cortex in depression: a randomized sham controlled study. J Affect Disord. 2011; 128(1-2):153–159.

52. Garcia-Toro M, Mayol A, Arnillas H, et al. Modest adjunctive benefit with transcranial magnetic stimulation in medication-resistant depression. J Affect Disord. 2001; 64(2-3):271–275.

53. Jhanwar VG, Bishnoi RJ, Jhanwar MR. Utility of repetitive transcranial stimulation as an augmenting treatment method in treatment-resistant depression. Indian J Psychol Med. 2011; 33(1):92–96.

54. Jhanwar VG, Bishnoi RJ, Singh L, et al. Utility of repetitive transcranial magnetic stimulation as an augmenting treatment method in treatment-resistant depression. Indian J Psychiatry. 2011; 53(2):145–148.

55. Verma R, Kumar N, Kumar S. Effectiveness of adjunctive repetitive transcranial magnetic stimulation in management of treatment-resistant depression: A retrospective analysis. Indian J Psychiatry. 2018; 60(3):329–333.

56. Conca A, Di Pauli J, Beraus W, et al. Combining high and low frequencies in rTMS antidepressive treatment: preliminary results. Hum Psychopharmacol. 2002; 17(7):353–356.

57. Qiao Y, Wang J, Zhu J, et al. Antidepressant Effect of Adjunct Repetitive Transcranial Magnetic Stimulation in Inpatients 60 Years and Older. J ECT. 2020; 36(3):216–221.

58. Zhang T, Sun W, Zhu J, et al. Effect of Adjunct Repetitive Transcranial Magnetic Stimulation in Elderly Patients with Acute Depressive Episode: Supporting Evidence from a Real-World Observation. Am J Geriatr Psychiatry. 2019; 27(1):91–92.

59. Mogg A, Pluck G, Eranti SV, et al. A randomized controlled trial with 4-month follow-up of adjunctive repetitive transcranial magnetic stimulation of the left prefrontal cortex for depression. Psychol Med. 2008; 38(3):323–333.

60. Rossini D, Lucca A, Zanardi R, et al. Transcranial magnetic stimulation in treatment-resistant depressed patients: a double-blind, placebo-controlled trial. Psychiatry Res. 2005; 137(1-2):1–10.

61. Sackeim HA. The definition and meaning of treatment-resistant depression. J Clin Psychiatry. 2001; 62 Suppl 16:10–17.

62. Smith JE, Peterchev AV. Electric field measurement of two commercial active/sham coils for transcranial magnetic stimulation. J Neural Eng. 2018; 15(5):054001.

63. Voigt JD, Leuchter AF, Carpenter LL. Theta burst stimulation for the acute treatment of major depressive disorder: A systematic review and meta-analysis. Transl Psychiatry. 2021; 11(1):330.

64. Cole EJ, Phillips AL, Bentzley BS, et al. Stanford Neuromodulation Therapy (SNT): A Double-Blind Randomized Controlled Trial. Am J Psychiatry. 2022; 179(2):132–141.

65. Cole EJ, Stimpson KH, Bentzley BS, et al. Stanford Accelerated Intelligent Neuromodulation Therapy for Treatment-Resistant Depression. Am J Psychiatry. 2020; 177(8):716–726.

66. Chandler GM, Iosifescu DV, Pollack MH, et al. RESEARCH: Validation of the Massachusetts General Hospital Antidepressant Treatment History Questionnaire (ATRQ). CNS Neurosci Ther. 2010; 16(5):322–325.

67. American Diabetes A. 9. Pharmacologic Approaches to Glycemic Treatment: Standards of Medical Care in Diabetes-2020. Diabetes Care. 2020; 43(Suppl 1):S98–S110.

68. Dunner DL, Aaronson ST, Sackeim HA, et al. A multisite, naturalistic, observational study of transcranial magnetic stimulation for patients with pharmacoresistant major depressive disorder: durability of benefit over a 1-year follow-up period. J Clin Psychiatry. 2014; 75(12):1394–1401.

69. Kedzior KK, Reitz SK, Azorina V, et al. Durability of the antidepressant effect of the high-frequency repetitive transcranial magnetic stimulation (rTMS) In the absence of maintenance treatment in major depression: a systematic review and meta-analysis of 16 double-blind, randomized, sham-controlled trials. Depress Anxiety. 2015; 32(3):193–203.

70. Senova S, Cotovio G, Pascual-Leone A, et al. Durability of antidepressant response to repetitive transcranial magnetic stimulation: Systematic review and meta-analysis. Brain Stimul. 2019; 12(1):119–128.

71. Rachid F. Maintenance repetitive transcranial magnetic stimulation (rTMS) for relapse prevention in with depression: A review. Psychiatry Res. 2018; 262:363–372.

72. Wilson S, Croarkin PE, Aaronson ST, et al. Systematic review of preservation TMS that includes continuation, maintenance, relapse-prevention, and rescue TMS. J Affect Disord. 2022; 296:79–88.

